# SARS-CoV-2 seroprevalence in Kaduna State, Nigeria during October/November 2021, following three waves of infection and immediately prior to detection of the Omicron variant

**DOI:** 10.1101/2021.12.21.21268166

**Authors:** Gloria D. Chechet, Jacob K.P. Kwaga, Joseph Yahaya, Annette MacLeod, Walt E. Adamson

**Affiliations:** Department of Biochemistry, Faculty of Life Sciences, Ahmadu Bello University, Zaria, Zaria, Nigeria; Africa Centre of Excellence for Neglected Tropical Diseases and Forensic Biotechnology, Ahmadu Bello University, Zaria, Nigeria; Department of Veterinary Public Health and Preventive Medicine, Faculty of Veterinary Medicine, Ahmadu Bello University, Zaria, Nigeria; Institute of Biodiversity, Life Sciences, and Animal Health, University of Glasgow, United Kingdom

## Abstract

Nigeria is the most populated country in Africa with an estimated ∼213 million inhabitants. As of November 2021 there have been three waves of SARS-CoV-2 infection in Nigeria but there has been only one seroprevalence survey conducted to assess the proportion of a population that have been infected, which was performed in December 2020 after the first wave of infection. To provide an update on seroprevalence in Nigeria, we conducted survey at one urban site (n=400) and one rural site (n=402) in Kaduna State, Nigeria during October and November 2021 following the third wave of infection. Seroprevalence for the urban and rural sites was 42.5% and 53.5% respectively (mean 48.0%). Symptoms associated with seropositivity were identified for each site. The overall seroprevalence among unvaccinated individuals was 45.4%. The data indicates an infection rate in Kaduna State at least 387 times greater than that derived from cases confirmed by PCR. Extrapolating to the whole of Nigeria, it would suggest there has been at least 96.7 million infections (compared to 206,138 confirmed cases at the time of surveillance). The work presented here will inform public health policy and deployment strategies for testing, treatment, and vaccination in Nigeria, and provide a baseline for SARS-CoV-2 seroprevalence in Nigeria immediately prior to the spread of the Omicron variant.

## Introduction

COVID-19 (caused by SARS-CoV-2 infection) was declared a pandemic by the World Health Organization (WHO) on 11 March 2020^1^. By 22 November 2021, 256,480,022 people across the world had confirmed infection, and 5,145,002 people had died as a result^2^.

Serological antibody tests to detect past exposure to SARS-CoV-2 are a valuable tool for assessing the proportion of a population that have been infected with the virus. They can detect evidence of infection from two weeks to several months following infection in both symptomatic and asymptomatic individuals. Identifying SARS-CoV-2 seroprevalence in a population has important implications for public health policy: determining the effectiveness of measures put in place to prevent the spread of the virus, providing evidence on whether further significant spread remains possible, and informing on deployment strategies for testing, treatment, and vaccination. Worldwide, as of 10 November 2021, 370 SARS-CoV-2 serological surveys had been published in peer-reviewed journals^3^. However, serological data for sub-Saharan Africa remains limited and often restricted to subgroups that might not represent the overall population such as healthcare workers. Since the beginning of the pandemic, several peer-reviewed SARS-CoV-2 seroprevalence studies have been conducted in sub-Saharan Africa^3^, however much of the data focuses on earlier stages of the pandemic. There have been just five peer-reviewed surveys published that describe data collected during 2021: all of which were in Eastern and Southern Africa^4,5,6,7,8^. Commentary in Lancet Global Health has called for SARS-CoV-2 seroprevalence studies to be carried out in sub-Saharan Africa ‘whenever possible’^9^.

Nigeria, in West Africa, is the continent’s most populated country with approximately 213 million inhabitants. As of November 2021, it had experienced three waves of SARS-CoV-2 infection (with peak infection rates occurring in July 2020, January 2021, and August 2021) (Figure 1)^2^. By 22 November 2021, official figures reported 213,589 confirmed cases and 2,974 deaths in Nigeria, however the per-capita testing rate is substantially lower than the African average (0.02 tests/capita in Nigeria compared with 0.06/capita continent-wide)^2^. In Nigeria there has been one peer-reviewed SARS-CoV-2 seroprevalence survey: examining samples collected in Anambra State during December 2020 (prior to the second wave of infection) and reporting an overall seroprevalence of 16.1%^10^. Since then, there have been a further 139.457 confirmed SARS-CoV-2 cases in Nigeria (65.3% of the country’s total confirmed cases)^2^. The country’s vaccination programme commenced in March 2021, and by 9 December 2021, 7.35 million inhabitants (3.48% of the population) had received at least one dose of a vaccine^2^. Earlier surveys in sub-Saharan Africa have highlighted differences in SARS-CoV-2 seroprevalence between urban and rural settings (with higher seroprevalence recorded in urban settings)^8^. To update the seroprevalence data for Nigeria following three waves of infection, we performed a seroprevalence survey at an urban and a rural site in Kaduna State, Nigeria during October and November 2021.

**Figure 1:**
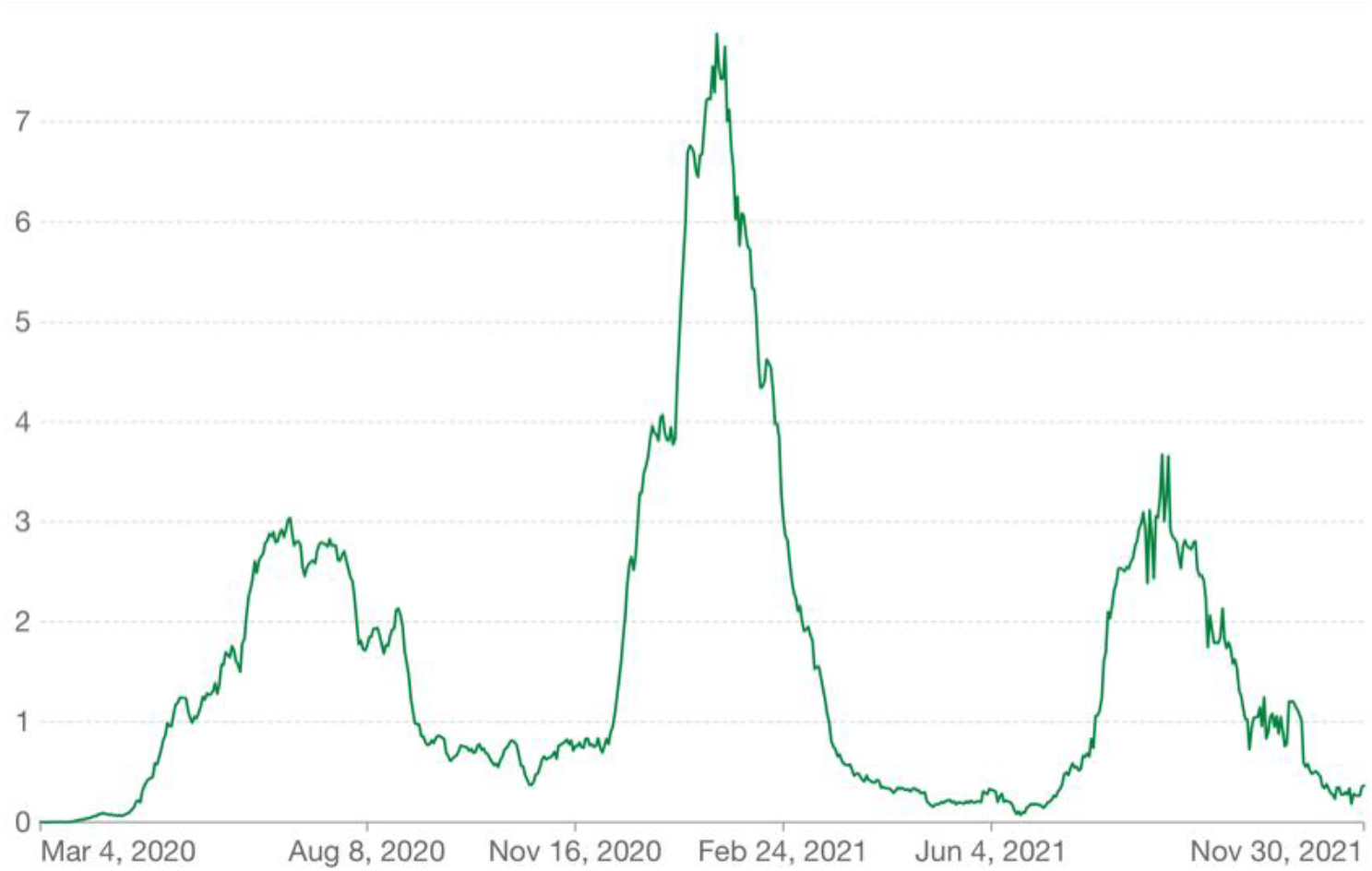
Daily new confirmed COVID-19 cases per million people in Nigeria, 7-day rolling average, 4 March 2020 to 30 November 2021. (Source: Our World In Data^2^.)

## Methods

Surveillance was carried out in accordance with the WHO’s Unity Studies Sero-Epidemiological Investigations Protocols^11^. We screened a total of 802 participants (Table 1) attending hospital outpatient units for reasons not related to COVID-19. Samples were collected at Yusuf Dantsoho Memorial Hospital, Tudun-Wada, Kaduna City (serving a predominantly urban population, n=400, sampling dates: 11 October 2021 to 5 November 2021) and Hajiya Gambo Suwaba General Hospital, Kofar-Gayan, Zaria (serving a predominantly rural population, n=402, sampling dates: 12 October 2021 to 8 November 2021). Informed consent was confirmed via signatures or thumb prints, while parental consent was obtained for participants who were less than 18 years old. Participants with learning disabilities were excluded from the study.

**Table 1:** Participant demographics: Kaduna City and Kofar-Gayan.

|  | Kaduna City | Kofar-Gayan |
| --- | --- | --- |
| Environment | Urban | Rural |
| Total | 400 | 402 |
| Female | 233 | 153 |
| Male | 167 | 248 |
| Sex not recorded | 0 | 1 |
| Age <21 | 130 | 91 |
| Age 21-30 | 78 | 226 |
| Age >30 | 159 | 82 |
| Age not recorded | 33 | 0 |
| Median age | 26 | 24 |
| Median household size | 7 | 12 |

Ethical approval for this study was obtained from Kaduna State Ministry of Health. Participants completed a questionnaire that captured age, sex, number of people living in their household, SARS-CoV-2 vaccination history, and 12-month history of symptoms previously associated with COVID-19 (aches and pains, diarrhoea, dry cough, fever, headache, loss of taste and/or smell, nausea, and sore throat^12^. Participants were also given the opportunity to indicate that they had not experienced any such symptoms in the previous 12 months.

Approximately 5 ml of venous blood was collected from each consenting participant. Tubes were left to clot at room temperature for one hour, then centrifuged at 2,500 rpm for 15 minutes. The resulting serum was analysed immediately. The CE-marked Biopanda COVID-19 IgM/IgG Rapid Test Kit was used to screen serum for antibodies, following the manufacturers’ instructions. Results were interpreted as described in Table 2. The presence of IgM antibodies indicates a recent infection (7-28 days prior to sampling), while IgG antibodies typically appear approximately 14 days after infection and endure for at least several months, providing some degree of immunity from further infection.

**Table 2:** interpretation of Biopanda COVID-19 IgM/IgG Rapid Test Kit results.

| Test result |  | Interpretation |
| --- | --- | --- |
| IgM | IgG |  |
| - | - | Seronegative |
| - | + | Seropositive, likely >28 days after infection |
| + | - | Seropositive, likely within 28 days of infection |
| + | + | Seropositive, likely 14-28 days after infection |

## Results

In total, 802 participants were screened for the presence of SARS-CoV-2 antibodies in Kaduna City (urban site, n=400) and Kofar-Gayan (rural site, n=402) during October and November 2021. Table 3 summarises antibody responses for Kaduna City and Kofar-Gayan.

**Table 3:** IgM and IgG seropositivity, Kaduna City and Kofar-Gayan. Z-tests were used to calculate p-values to compare subgroups.

|  | Kaduna City (n=400) | Kofar-Gayan (n=402) | p-value |
| --- | --- | --- | --- |
| IgG positive only | 129 (32.25%) | 189 (47.01%) | <0.001 |
| IgM positive only | 23 (5.75%) | 13 (3.23%) | 0.085 |
| IgG and IgM positive | 18 (4.50%) | 13 (3.23%) | 0.352 |
| All IgG positive | 147 (36.75%) | 202 (50.25%) | <0.001 |
| All IgM positive | 41 (10.25%) | 26 (6.47%) | 0.052 |
| All positive | 170 (42.50%) | 215 (53.48%) | 0.002 |
| Negative | 230 (57.50%) | 187 (46.52%) | 0.002 |

### SARS-CoV-2 seroprevalence data for Kaduna City (urban site)

In Kaduna City, antibodies were detected in 170 participants (42.5%) of whom 41 (10.25% of all Kaduna City participants) were IgM positive, suggestive of a recent infection. No detectable difference between seroprevalence rates in female and male participants was observed (p=0.064) (Table 4). Seroprevalence among participants who were under 21 years of age was lower (p<0.00001) than older age groups (Table 5), and the average age of seropositive participants (33.1 years) was greater (p=0.018) than those who were seronegative (28.7 years). No significant differences were observed between SARS-CoV-2 seropositive and seronegative participants in terms of household size. Vaccination against SARS-CoV-2 was reported in 33 participants (8.25%), 30 of whom reported receiving the AZD1222 (ChAdOx1 nCoV-19) vaccine (manufactured by AstraZeneca) while 3 reported having been vaccinated with Ad26.COV2.S (manufactured by Janssen).

**Table 4:** Seropositivity in females and males Kaduna City and Kofar-Gayan. Z-tests were used to calculate p-values to compare subgroups.

|  | Females | Males | p-value (identifying difference between sexes) |
| --- | --- | --- | --- |
| Kaduna City (urban) | 90/233 (38.6%) | 80/167 (47.9%) | 0.064 |
| Kofar-Gayan (rural) | 80/153 (52.3%) | 134/247 (54.3%) | 0.704 |
| p-value (identifying difference between locations) | 0.008 | 0.204 |  |

**Table 5:** Seropositivity by age group, Kaduna City and Kofar-Gayan. Confidence intervals were calculated as ±1.96 x standard error of the proportion who are seropositive.

| Age group | Kaduna City |  | Kofar-Gayan |  |
| --- | --- | --- | --- | --- |
|  | % positive | 95% CI | % positive | 95% CI |
| Under 21 | 30.7 | 22.8% - 38.6% | 48.3 | 38.0% - 58.6% |
| 21 – 30 | 60.3 | 49.4% - 71.2% | 55.3 | 48.8% - 61.8% |
| Over 30 | 50.9 | 43.1% - 58.7% | 52.4 | 41.6% - 63.2% |

### Symptoms (Kaduna City)

Participants reported experiencing an average of 1.6 of the 8 common COVID-19 symptoms listed in the questionnaire. Aches and pains, dry cough, fever, loss of taste and/or smell, nausea, and sore throat were associated with SARS-CoV-2 seropositivity (Table 6). SARS-Cov-2 seropositive participants reported more symptoms than those who were seronegative (p<0.001), and reporting no symptoms was associated with being seronegative.

**Table 6:** SARS-CoV-2 antibody test results by symptoms reported, Kaduna City. Symptoms associated with seropositivity are highlighted in bold. P-values were calculated using a chi-squared test.

| Symptom (total number reporting) | SARS-CoV-2 seropositive (%) | Seropositive OR (95% CI) | p-value |
| --- | --- | --- | --- |
| **Aches and pains (74)** | **57 (77.0%)** | **6.3 (3.5 - 11.4)** | **<0.001** |
| Diarrhoea (24) | 9 (37.5%) | 0.9 (0.4 – 2.2) | 0.877 |
| **Dry cough (67)** | **43 (64.2%)** | **2.9 (1.7 – 5.0)** | **<0.001** |
| **Fever (112)** | **72 (64.3%)** | **2.0 (1.2 – 3.2)** | **<0.001** |
| Headache (221) | 95 (43.0%) | 1.0 (0.7 – 1.6) | 0.827 |
| **Loss of taste and/or smell (72)** | **63 (87.5%)** | **14.4 (6.9 – 30.2)** | **<0.001** |
| **Nausea (22)** | **15 (68.1%)** | **3.1 (1.2 – 7.7)** | **0.012** |
| **Sore throat (62)** | **53 (85.5%)** | **11.1 (5.3 – 23.3)** | **<0.001** |
| No symptoms reported (68) | 16 (23.5%) | 0.4 (0.2 – 0.6) | <0.001 |

### SARS-CoV-2 seroprevalence data for Kofar-Gayan (rural site)

In Kofar-Gayan, antibodies were detected in 215 participants (53.5%), of whom 26 (6.5% of all Kofar-Gayan participants) were IgM seropositive (Table 3). No detectable differences in seroprevalence rates were detected when comparing female and male participants (Table 4), age groups (Table 5), or household sizes. Vaccination against SARS-CoV-2 was reported in 16 participants (4.0%), all of whom reported receiving the AZD1222 (ChAdOx1 nCoV-19) vaccine.

### Symptoms (Kofar-Gayan)

Participants reported experiencing an average of 3.7 of the common COVID-19 symptoms listed in the questionnaire. Loss of taste and/or smell and sore throat were associated with seropositivity (Table 7). Seropositive participants reported experiencing more symptoms than those who were seronegative (p=0.010).

**Table 7:** SARS-CoV-2 antibody test results by symptoms reported, Kofar-Gayan. Symptoms associated with seropositivity are highlighted in bold. P-values were calculated using a chi-squared test.

| Symptom (total number reporting) | SARS-CoV-2 seropositive (%) | Seropositive OR (95% CI) | p-value |
| --- | --- | --- | --- |
| Aches and pains (365) | 199 | 1.6 (0.8 – 3.1) | 0.190 |
| Diarrhoea (33) | 16 | 0.8 (0.4 – 1.6) | 0.548 |
| Dry cough (157) | 86 | 1.1 (0.7 – 1.6) | 0.677 |
| Fever (333) | 172 | 0.6 (0.4 – 1.1) | 0.101 |
| Headache (373) | 204 | 2.0 (0.9 – 4.3) | 0.081 |
| **Loss of taste and/or smell (62)** | **50** | **4.4 (2.3 – 8.6)** | **<0.001** |
| Nausea (137) | 74 | 1.0 (0.7 – 1.6) | 0.878 |
| **Sore throat (46)** | **31** | **1.9 (1.0 – 3.7)** | **0.044** |
| No symptoms reported (14) | 6 | 0.6 (0.2 – 1.9) | 0.417 |

### Comparison of SARS-CoV-2 seroprevalence data for Kaduna City and Kofar-Gayan

Overall seroprevalence was higher in Kofar-Gayan than Kaduna City (p=0.002). No difference in seroprevalence rates was detected in male participants at the two sites, however seroprevalence rates in females in Kaduna City were lower than in Kofar-Gayan (p=0.008) (Table 5). Although IgM antibodies were detected in a higher percentage of participants in Kaduna City (Table 4), this difference was not significant (p=0.055). Participants in Kaduna were more likely to have been vaccinated against SARS-CoV-2 (p=0.011). Participants in Kofar-Gayan reported experiencing more symptoms associated with COVID-19 (p<0.001).

## Discussion

Serological data for SARS-CoV-2 infection in sub-Saharan Africa is limited. To date, one previous peer-reviewed serological survey has taken place in Nigeria^10^, and the country has subsequently experienced two further waves of SARS-CoV-2 infection^2^. Although the surveillance described here was limited to two sites, it provides evidence on the proportion of the Nigerian population who have been exposed to the virus. The surveillance method deployed does not, however, differentiate between seropositivity as a result of infection or as a result of vaccination. Other caveats include those individuals who are unable to mount a detectable antibody response, and the fact that SARS-CoV-2 antibody titres have been shown to decline in the months following exposure. Due to the delay between exposure and development of SARS-CoV-2 antibodies, it is likely the seropositive participants of this survey will have been infected by or vaccinated against the virus by 27 September 2021 (two weeks prior to the commencement of sampling).

Across the two sites examined in this study, the overall seroprevalence was 48.0%, and the seroprevalence among unvaccinated participants was 45.4%. If the seroprevalence data reported here is representative of Kaduna State (population 8.25 million^13^), it would indicate at least 3.75 million SARS-CoV-2 infections since the pandemic began: a figure 387 times greater than the state’s 9,695 confirmed cases as of 27 September 2021^13^. Extrapolating to the whole of Nigeria (population 213 million), this would suggest at least 96.7 million infections. This is in stark contrast to the number of confirmed cases at that time: 206,138^13^.

During sample and data collection, the proportion of the Nigerian population who had received at least one dose of a SARS-CoV-2 vaccine was between 2.3% and 2.7%^2^. Self-reporting of vaccination in this survey indicated that 8.25% of participants in Kaduna City and 4.0% of participants in Kofar-Gayan had received at least one dose of a vaccine. This indicates that participants were not representative of the overall Nigerian population in terms of vaccination status. The factors responsible are not clear, although in Nigeria older age groups were prioritised for SARS-CoV-2 vaccination^14^, and the median age of participants in this survey (25) was older than the reported median age for the Nigerian population (18)^15^. The observed differences in vaccination rates between Kaduna City and Kofar-Gayan suggest potentially uneven distribution of vaccines between urban and rural populations in Nigeria.

In contrast to previous seroprevalence data comparing urban and rural areas^8^, SARS-CoV-2 antibody seroprevalence was higher in Kofar-Gayan than in Kaduna City. Variation between geographical locations in one country is a common feature of the serological surveillance of respiratory viruses^16^, with multiple factors promoting these differences. One possible factor is household size. Larger household sizes have previously been implicated in the spread of the virus^17^. Participants in Kofar-Gayan has a median household size of 12 compared to those in Kaduna City (7), suggesting a possible explanation for this variation. Within Kaduna City, the lower seroprevalence observed in females and younger age groups highlights population subgroups who, without protective antibodies, may be susceptible to future waves of SARS-CoV-2 infection, and who might benefit from targeted public health strategies.

Symptoms identified as being associated with COVID-19 reported here are consistent with previous studies. Participants in Kofar-Gayan reported more symptoms than those in Kaduna City: an observation that could reflect an increased circulation of other pathogens, or other socioeconomic factors.

On 26 November 2021, WHO classified SARS-CoV-2 variant B.1.1.529 a variant of concern, and designated it with the name Omicron^18^. Preliminary data indicates that Omicron is likely to replace Delta as the world’s predominant SARS-CoV-2 variant^19^: a change that would likely result in increased rates of SARS-CoV-2 infection. Obtaining baseline measurements that record the proportion of the population with some antibody response to previous variants of SARS-CoV-2 immediately prior to the introduction of Omicron will inform intervention strategies.

## Conclusions

This study provides SARS-CoV-2 seroprevalence data for the most populated sub-Saharan African country. It provides detailed information related to seroprevalence for two locations in Kaduna State. Further studies are required to ascertain whether the seroprevalence reported is consistent across the country, and to record longitudinal seroprevalence changes. It indicates infection rates in Nigeria might be at least 387 times greater than figures for confirmed cases, and provides evidence that as of October/November 2021, 48% of the Nigerian population have been exposed to the virus via infection or vaccination, and that approximately 52% did not carry SARS-CoV-2 antibodies. The work presented here will inform public health policy and deployment strategies for testing, treatment, and vaccination in Nigeria, and provide a record of SARS-CoV-2 seroprevalence in Nigeria during October/November 2021, following three waves of infection. Additionally, it provides a baseline for SARS-CoV-2 seroprevalence in Nigeria prior to the spread of the Omicron variant.

## Data Availability

All data produced in the present study are available upon reasonable request to the authors.

